# Burden of Bronchiectasis Among COPD Patients in Bangladesh: Insights from a Cross-Sectional Study

**DOI:** 10.64898/2025.12.16.25342432

**Authors:** Imran Hossain, Marzia Sultana Shanta, Kazi Hataf Rahbor Abrar, Sohel Rana, Sabrina Rahman Tarafder, Md Mazidul Haque

## Abstract

**Background:** Bronchiectasis is an increasingly recognized structural complication among patients with chronic obstructive pulmonary disease (COPD), yet evidence from Bangladesh remains limited. This study aimed to determine the prevalence, clinical characteristics, radiological patterns, and associated factors of bronchiectasis among COPD patients in tertiary-level hospitals.

**Methods:** A cross-sectional study was conducted among 129 COPD patients regardless of age distribution, all are met GOLD criteria and underwent high-resolution computed tomography (HRCT). Data on sociodemographic, behavioral, clinical, radiological, laboratory, and quality-of-life characteristics were analyzed using descriptive statistics, chi-square tests, t-tests, correlation analyses, and binary logistic regression.

**Results:** Bronchiectasis was detected in 73.6% of COPD patients, with cylindrical bronchiectasis being the most common subtype (48.4%). Patients with bronchiectasis had a significantly longer duration of COPD (7.58 ± 3.36 years vs. 2.51 ± 1.67 years; p < 0.001) and more frequent symptoms, including chronic cough (72.1%), purulent sputum (34.9%), and higher dyspnea grades. Mucus plugging showed a perfect association with bronchiectasis (p = 0.001). Significant predictors of bronchiectasis included rural residence (AOR 5.82; 95% CI 1.34–25.29) and smoking habit (AOR 3.69; 95% CI 1.01–13.49). A weak but significant negative correlation was found between serum albumin and CRP (r = −0.211; p = 0.016), indicating systemic inflammation, while smoking duration was negatively correlated with FEV□ (r = −0.174; p = 0.048). Quality of life was markedly impaired, with over 70% reporting poor or fair status.

**Conclusion:** Bronchiectasis is highly prevalent among COPD patients in Bangladesh and is associated with longer disease duration, greater symptom burden, functional impairment, structural lung abnormalities, and poor quality of life. Rural residence, smoking, and mucus plugging emerged as key determinants. Early HRCT-based screening, phenotype-specific management, reduction of biomass and tobacco exposure, and improved rural respiratory care are essential to mitigate disease progression and improve outcomes.

## Introduction

Chronic obstructive pulmonary disease (COPD) represents one of the most significant global health challenges of the 21st century. It is a progressive, debilitating respiratory disorder characterized by persistent airflow limitation, chronic airway inflammation, and recurrent exacerbations that undermine quality of life and increase the risk of premature death. The Global Initiative for Chronic Obstructive Lung Disease (GOLD) identifies COPD as a major cause of morbidity and mortality worldwide, with more than 3 million deaths annually—approximately 80% of which occur in low- and middle-income countries where health systems are often under-resourced and exposures to environmental and occupational pollutants are high. COPD is now widely recognized as a heterogeneous condition encompassing a spectrum of phenotypes, clinical patterns, and structural abnormalities that vary widely between populations.

Among the emerging phenotypes within COPD, bronchiectasis has gained considerable attention for its strong clinical relevance. Bronchiectasis is characterized by permanent dilatation of the bronchi due to chronic inflammation, impaired muco-ciliary clearance, and repeated infections leading to progressive airway damage. Historically, bronchiectasis was considered a separate disease entity; however, contemporary research suggests that it frequently coexists with COPD and may represent a distinct “overlap phenotype” with specific clinical, radiological, and microbiological characteristics. Systematic reviews show that the prevalence of bronchiectasis among COPD patients ranges from 25% to nearly 70%, depending on study populations, disease severity, and diagnostic criteria^1,2^. The increasing detection of bronchiectasis in COPD is partly due to wider availability of high-resolution computed tomography (HRCT), which has revolutionized our understanding of airway structural abnormalities.

The coexistence of COPD and bronchiectasis is not merely additive; rather, it gives rise to a synergistic and clinically more severe phenotype. Patients with this overlap frequently exhibit more intense airway inflammation, worse airflow limitation, and significantly greater symptom burden, especially chronic sputum production and persistent cough^3,4^. Recurrent bacterial infections—most notably due to *Pseudomonas aeruginosa* and other potentially pathogenic microorganisms—are substantially more common in this phenotype and contribute to a vicious cycle of infection, inflammation, and further airway destruction^1,4^. These microbiological features are strongly associated with more frequent acute exacerbations, prolonged recovery periods, and greater risk of hospitalization. Moreover, studies have consistently demonstrated that patients with COPD–bronchiectasis overlap have higher systemic inflammatory markers, suggesting a more aggressive underlying disease mechanism^2,5^.

Radiological studies further support the significance of this overlap. In emphysema-predominant COPD—the form most commonly associated with smoking—structural airway changes such as bronchiectasis are also highly prevalent, highlighting the shared pathophysiological pathways between emphysema and bronchiectasis, including chronic inflammation, protease–antiprotease imbalance, tissue destruction, and impaired mucociliary clearance^5^. These shared pathways may explain why bronchiectasis often appears in advanced stages of COPD and may serve as a marker of severe disease progression. Clinically, the overlap phenotype is associated with greater mortality, increased hospitalizations, and poorer long-term outcomes compared to COPD alone^2,3,6^.

The epidemiology of bronchiectasis is changing worldwide, with several countries reporting rising incidence and prevalence. A population-based study from Italy showed a steady increase in bronchiectasis diagnoses over time, particularly among the elderly^7^. Similar trends have been observed in Europe, Asia, and North America. This rising trend reflects both true increases in disease burden and better diagnostic practices. However, substantial underdiagnosis persists globally, especially in low-resource settings where HRCT is not widely available. This under-recognition contributes to delayed management, inappropriate treatment, and increased morbidity.

The South Asian region—including Bangladesh—carries a particularly heavy burden of chronic respiratory diseases. COPD prevalence is high, driven by tobacco use, biomass fuel exposure, indoor air pollution, occupational hazards, and a considerable burden of pulmonary infections such as tuberculosis^8^. In Bangladesh, nearly 12–13% of adults are estimated to have COPD, with prevalence reaching 20–40% in certain high-risk occupations and rural populations^8^. This epidemiological landscape creates an ideal environment for the development of bronchiectasis, especially post-infectious forms. Tuberculosis scarring and untreated lower respiratory infections continue to be important contributors to structural lung disease, including bronchiectasis.

Local studies from Bangladesh reveal that bronchiectasis patients often present with neutrophil-dominant inflammatory phenotypes associated with severe radiological involvement and frequent exacerbations^9^. Nutritional and systemic inflammatory markers—such as serum albumin—have been shown to correlate with bronchiectasis severity and may serve as accessible, low-cost prognostic tools in resource-limited settings^10^. Furthermore, research indicates that COPD significantly reduces health-related quality of life, and this impairment becomes more pronounced in the presence of coexisting bronchiectasis^11^. The overlap phenotype often leads to greater functional limitations, increased dyspnoea, reduced exercise tolerance, and a greater psychological burden.

The complexity of COPD–bronchiectasis overlap presents unique therapeutic challenges. Standard COPD treatments—such as long-acting bronchodilators and inhaled corticosteroids—may be insufficient or even inappropriate for some overlap patients. Inhaled corticosteroids, while beneficial for eosinophilic COPD, may increase the risk of infections in bronchiectasis-dominant phenotypes. Systemic corticosteroids may offer short-term benefit during acute bronchiectasis exacerbations^12^, but long-term use remains controversial. Adjunct therapies, including long-term macrolides, mucolytics, airway clearance techniques, and targeted antibiotic regimens, are often required in bronchiectasis but may be overlooked when bronchiectasis is not recognized. Older oral bronchodilators like theophylline still play a role in some settings but have limitations due to toxicity; newer agents such as doxofylline demonstrate improved safety with comparable efficacy and may be particularly useful in resource-constrained environments^13^.

Despite the global recognition of the COPD–bronchiectasis overlap, data from Bangladesh are scarce. Limited access to HRCT, underdeveloped respiratory diagnostic pathways, and low clinical suspicion contribute to underdiagnosis. As a result, many patients with bronchiectasis may be misclassified as having severe COPD, leading to incomplete management and worse outcomes. Given Bangladesh’s unique epidemiological profile—high smoking rates, extensive biomass exposure, and widespread post-infectious lung disease—understanding the extent and pattern of bronchiectasis among COPD patients is critical for improving clinical care and public health planning.

This thesis is designed to fill this knowledge gap by comprehensively examining the clinical, epidemiological, and radiological characteristics of bronchiectasis among patients with COPD. By identifying associated factors and disease patterns, this research aims to support early diagnosis, phenotype-based treatment strategies, and improved patient outcomes in the Bangladeshi context^14^. Ultimately, strengthening local evidence on COPD–bronchiectasis overlap will contribute to more effective healthcare policy, targeted interventions, and better allocation of limited resources within the country’s respiratory health services.

## Methodology

This cross-sectional descriptive study was conducted to comprehensively determine the prevalence, phenotypic distribution, and clinical burden of bronchiectasis among patients with chronic obstructive pulmonary disease (COPD) in Bangladesh. The study was carried out across multiple tertiary-level hospitals with advanced diagnostic infrastructure, including spirometry and high-resolution computed tomography (HRCT), ensuring rigorous, standardized, and reliable assessment of all participants. The multi-center design allowed for a heterogeneous patient population, enhancing the generalizability of findings to the broader COPD population in Bangladesh.

Eligible participants were recruited using consecutive sampling, targeting adults aged 40 years and above who presented to either outpatient or inpatient departments during the study period. Inclusion required a confirmed diagnosis of COPD according to the Global Initiative for Chronic Obstructive Lung Disease (GOLD) criteria, defined as a post-bronchodilator FEV /FVC ratio of less than 0.70. Only patients who had undergone HRCT imaging within the study timeframe and who provided written informed consent were enrolled. Exclusion criteria were carefully defined to reduce diagnostic ambiguity and confounding, including patients with active pulmonary tuberculosis or a history of tuberculosis in the preceding 12 months, bronchial asthma, lung malignancy, cystic fibrosis, other structural lung diseases, or those unable or unwilling to participate. These criteria ensured that bronchiectatic changes observed were attributable specifically to COPD rather than other pulmonary pathologies.

Sample size estimation was conducted using the standard formula for a single population proportion, accounting for a 10% non-response rate. This yielded a target sample of 129 participants, surpassing the minimum requirement of 118 to maintain adequate statistical power. Purposive recruitment of all eligible patients ensured that the study achieved this target, capturing a representative spectrum of COPD severity and phenotypic presentations.

Data collection was performed using a structured, interviewer-administered questionnaire developed after an extensive literature review and expert consultation to ensure content validity. The questionnaire encompassed comprehensive socio-demographic variables (age, sex, occupation, education level, socioeconomic status, and place of residence) and detailed behavioral and environmental exposures, including smoking history, duration and intensity of smoking, biomass fuel exposure, and occupational contact with respiratory irritants. Clinical variables included COPD duration, symptom profile, frequency of exacerbations in the preceding year, sputum characteristics, history of hospital admissions, and comorbidities such as hypertension, diabetes mellitus, and cardiovascular disease.

Radiological evaluation was performed using HRCT scans conducted according to standardized protocols. Each scan was interpreted independently by an experienced consultant radiologist blinded to the clinical information to minimize observer bias. Bronchiectasis was diagnosed based on established HRCT criteria, including a broncho-arterial ratio exceeding 1.0, lack of normal bronchial tapering, and the presence of associated features such as mucus plugging, bronchial wall thickening, and lobar involvement. Detailed documentation included the anatomical distribution, pattern of lobe involvement, severity, and classification into neutrophilic, eosinophilic, or non-neutrophilic phenotypes, based on internationally recognized criteria.

Laboratory data, including serum albumin and total leukocyte count, were retrieved from hospital records. Hypoalbuminemia was defined as serum albumin <3.5 g/dL. Disease severity for bronchiectasis was assessed using the Bronchiectasis Severity Index (BSI), while COPD severity was classified according to GOLD staging based on post-bronchodilator FEV percent predicted. Health-related quality of life (HRQoL) was assessed using the St. George’s Respiratory Questionnaire (SGRQ), a validated instrument that evaluates symptoms, physical limitations, and psychosocial impact. Higher SGRQ scores indicated greater impairment. For analytic purposes, COPD was defined as persistent airflow limitation (FEV /FVC <0.70), bronchiectasis was defined radiologically using HRCT, and co-occurrence of both conditions was classified as bronchiectasis–COPD overlap syndrome (BCOS). Patients experiencing two or more exacerbations or at least one hospitalization in the preceding 12 months were categorized as frequent exacerbators, reflecting higher clinical burden.

Data quality was ensured through meticulous review for completeness, coding, and entry into SPSS version 26.0. Descriptive statistics, including frequency distributions, percentages, means, and standard deviations, summarized participant characteristics. Inferential analyses employed Chi-square or Fisher’s exact tests for categorical variables and independent sample t-tests or one-way ANOVA for continuous variables, with normality assessments guiding appropriate test selection. Variables demonstrating statistical significance in bivariate analyses or clinical relevance were incorporated into binary logistic regression models to identify independent predictors of bronchiectasis among COPD patients. A two-tailed p-value <0.05 was considered statistically significant.

Ethical rigor was maintained in accordance with the Declaration of Helsinki (2013). All participants were fully informed about the study’s objectives, potential risks and benefits, and their rights to voluntary participation and withdrawal without repercussions to their clinical care. Written informed consent was obtained, and confidentiality was ensured via unique participant identifiers and restricted access to data, available only to the principal investigator. The study was designed with the overarching goal of generating robust epidemiological evidence to inform improved diagnostic strategies, individualized clinical management, and public health interventions for COPD and bronchiectasis in Bangladesh. By systematically characterizing the clinical and radiological spectrum of BCOS, this study aimed to fill critical knowledge gaps and provide a foundation for future longitudinal and interventional research in this population.

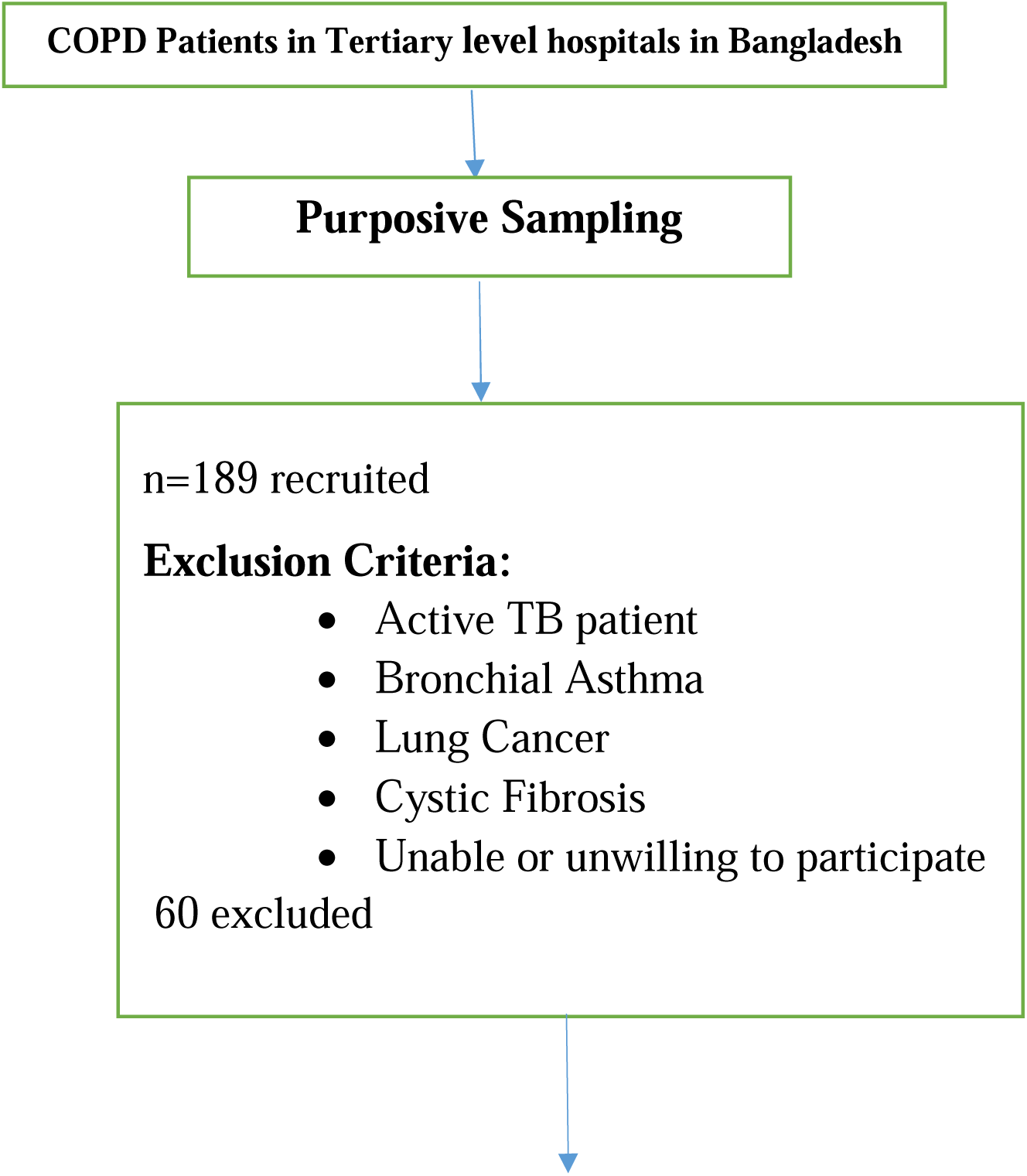
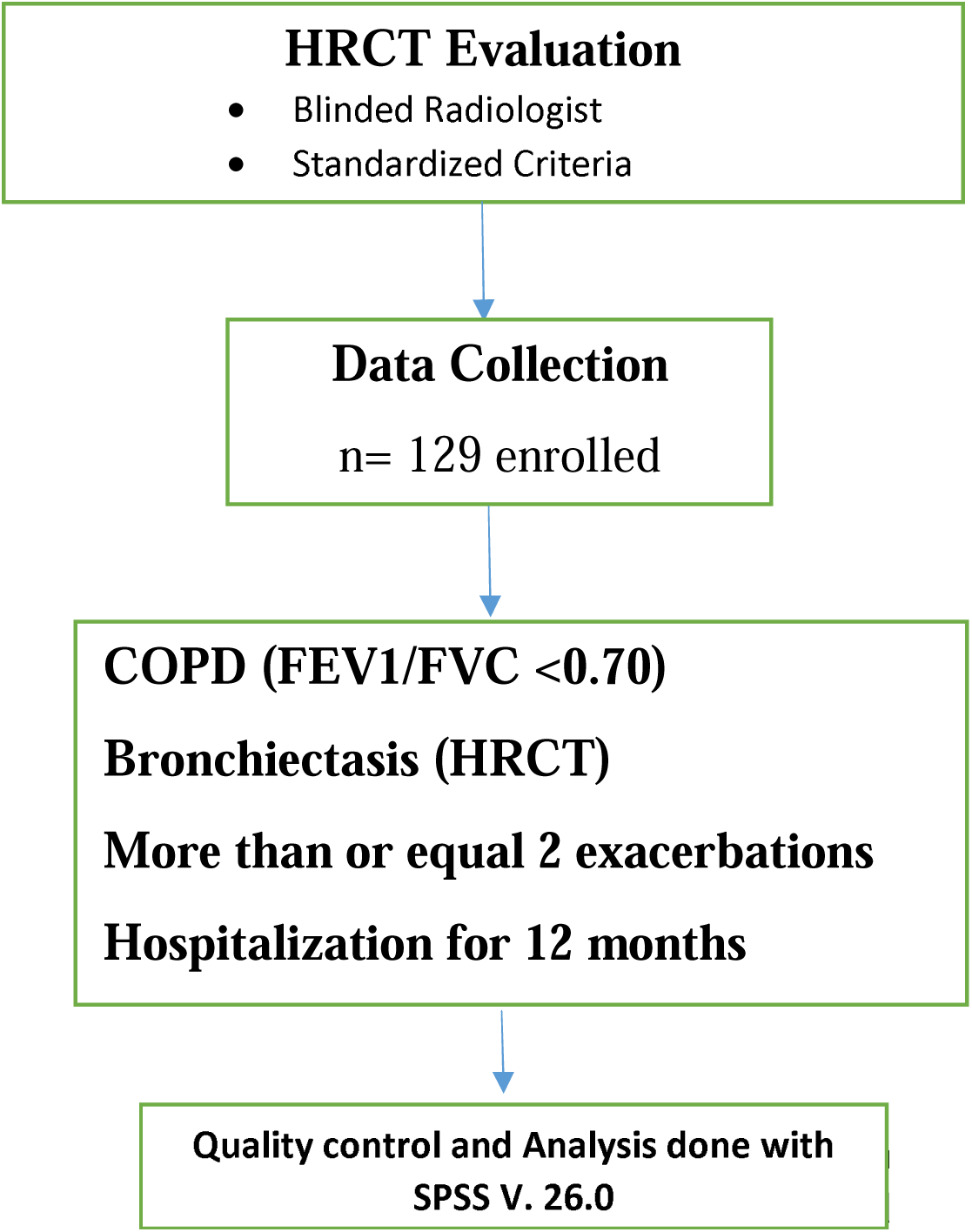

## RESULTS

This cross-sectional study assessed the burden of bronchiectasis among 129 patients with COPD in Bangladesh. Data were analyzed using SPSS version 26 and are presented through descriptive statistics, comparative analyses, correlations, and regression modeling. The findings are organized into socio-demographic characteristics, behavioral and personal factors, clinical and disease profiles, respiratory symptoms and functional status, radiological patterns, laboratory and pulmonary function results, quality of life, and associations between key variables.

Table 1 summarizes the socio-demographic profile of the 129 respondents. Most were middle-aged to elderly, with 45.0% aged 51–60 years and 28.7% aged 40–50 years; only 5.4% were below 40. Males constituted 55.0% of the sample, indicating a slightly higher male participation.

**Table 1.** Socio demographic characteristics of the respondents (n= 129)

| Variables | Category | Frequency | Percent(%) |
| --- | --- | --- | --- |
| Age | <40 | 7 | 5.4 |
|  | 40-50 | 37 | 28.7 |
|  | 51-60 | 58 | 45 |
|  | >60 | 27 | 20.9 |
| Gender | Male | 71 | 55 |
|  | Female | 58 | 45 |
| Religion | Islam | 123 | 95.42 |
|  | Hindu | 5 | 3.78 |
|  | Buddhist | 1 | 0.8 |
| Marital Status | Married | 100 | 77.5 |
|  | Unmarried | 11 | 8.5 |
|  | Widow/Widowed | 14 | 10.9 |
|  | Divorced | 4 | 3.1 |
| Educational status | Primary | 41 | 31.7 |
|  | Secondary & higher secondary | 28 | 21.7 |
|  | Graduation and above | 26 | 20.2 |
|  | No formal Education | 34 | 26.4 |
| Income of Respondents | <10000 | 4 | 3.1 |
|  | 10000-20000 | 20 | 15.5 |
|  | 20001-30000 | 53 | 41.1 |
|  | 30001-40000 | 20 | 15.5 |
|  | >40000 | 5 | 3.9 |
|  | No income | 27 | 20.9 |
| Occupation of respondents | Business | 4 | 3.2 |
|  | Doctor | 3 | 2.3 |
|  | Driver | 11 | 8.5 |
|  | Farmer | 12 | 9.3 |
|  | Garment Worker | 16 | 12.4 |
|  | Housewife | 31 | 24 |
|  | Office worker | 14 | 10.9 |
|  | Retired | 13 | 10.1 |
|  | Shopkeeper | 15 | 11.6 |
|  | Teacher | 2 | 1.6 |
|  | Unemployed | 8 | 6.2 |
| Residence of respondents | Urban | 92 | 71.3 |
|  | Rural | 37 | 28.7 |

The overwhelming majority were Muslim (95.42%), reflecting the general demographic distribution of Bangladesh. Most respondents were married (77.5%), with smaller proportions widowed (10.9%), unmarried (8.5%), and divorced (3.1%).

In terms of education, 31.7% had completed primary education, 21.7% secondary or higher secondary, and 20.2% held graduate or higher qualifications, while 26.4% had no formal education.

Monthly income varied, with 41.1% earning 20,001–30,000 BDT. One-fifth (20.9%) had no income, likely reflecting dependents such as housewives or retirees. Occupational distribution was diverse: 24.0% were housewives, followed by garment workers (12.4%), shopkeepers (11.6%), office workers (10.9%), farmers (9.3%), and several other professions. A majority (71.3%) resided in urban areas.

Overall, the socio-demographic profile reflects a mixed group of predominantly middle-aged, married, urban residents with moderate educational and economic backgrounds.

Table 2 shows that 64.3% of respondents had never smoked, while 27.1% were current smokers and 8.5% former smokers. Among those who smoked, the mean duration was 10.08 ± 9.87 years and the average number of cigarettes smoked per day was 6.45 ± 9.39.

**Table 2:**
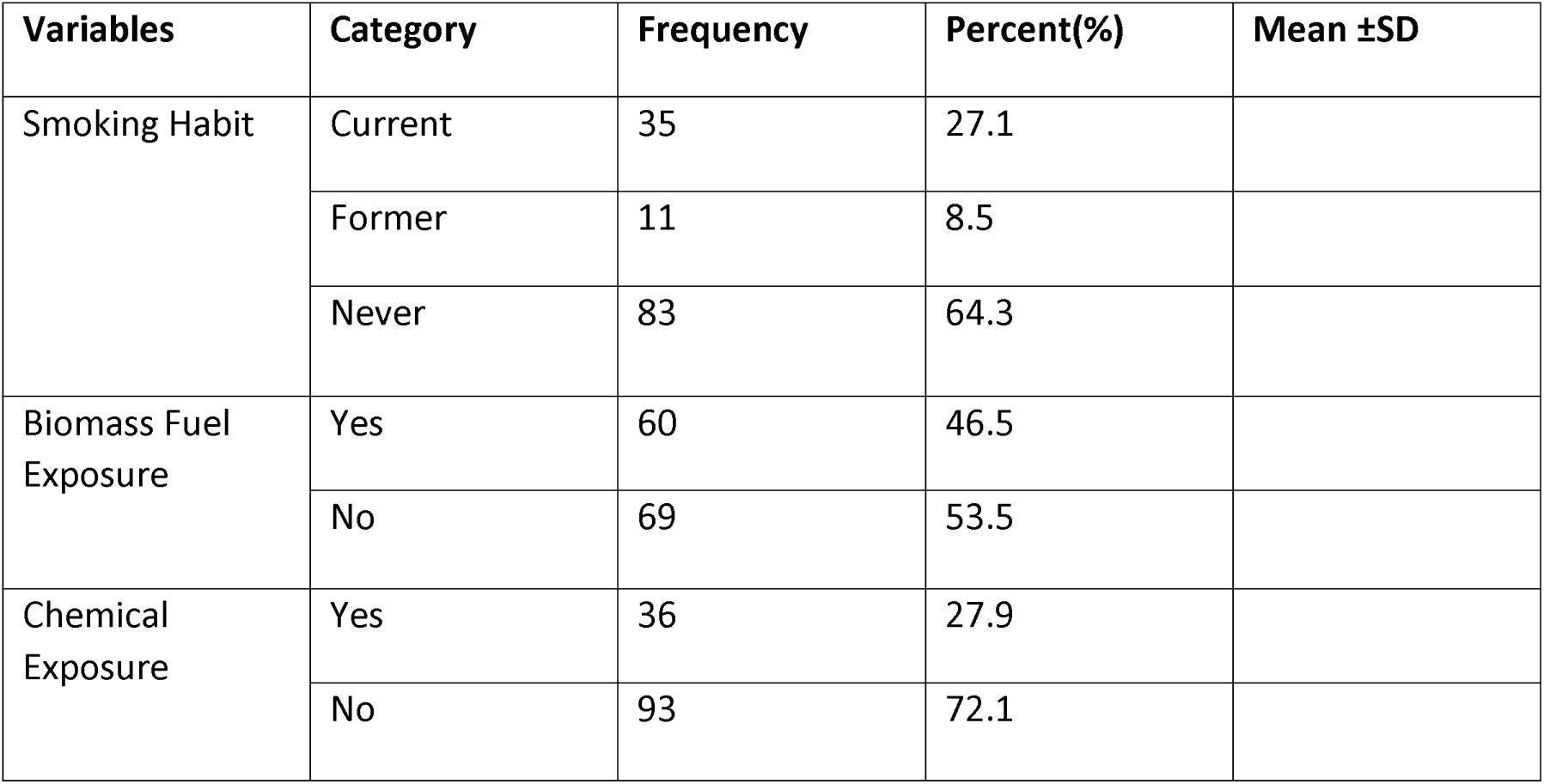
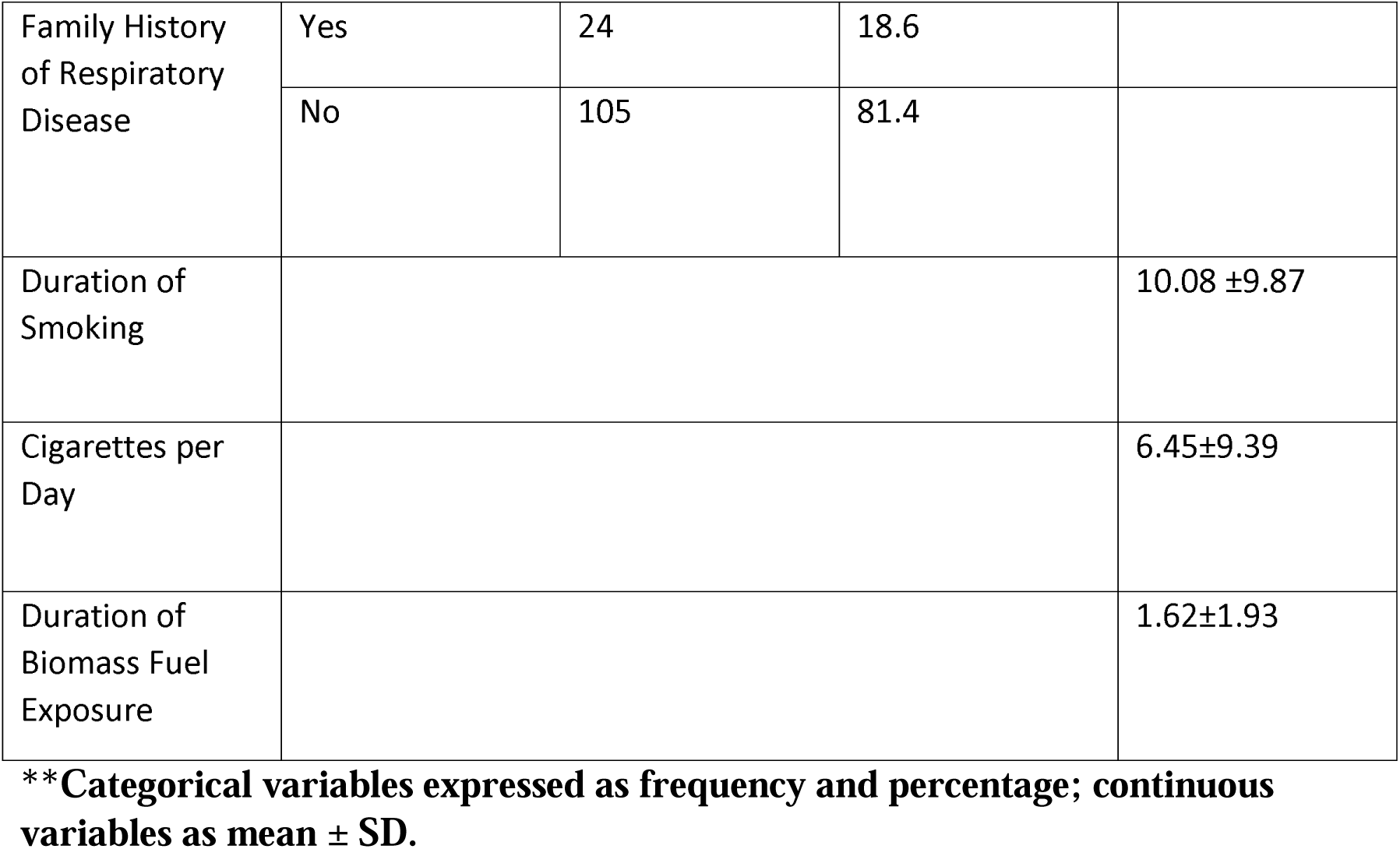
Distribution of respondents by personal and behavioral characteristics (n = 129)

| Variables | Category | Frequency | Percent(%) | Mean ±SD |
| --- | --- | --- | --- | --- |
| Smoking Habit | Current | 35 | 27.1 |  |
|  | Former | 11 | 8.5 |  |
|  | Never | 83 | 64.3 |  |
| Biomass Fuel Exposure | Yes | 60 | 46.5 |  |
|  | No | 69 | 53.5 |  |
| Chemical Exposure | Yes | 36 | 27.9 |  |
|  | No | 93 | 72.1 |  |
| Family History of Respiratory Disease | Yes | 24 | 18.6 |  |
|  | No | 105 | 81.4 |  |
| Duration of Smoking |  |  |  | 10.08 ±9.87 |
| Cigarettes per Day |  |  |  | 6.45±9.39 |
| Duration of Biomass Fuel Exposure |  |  |  | 1.62±1.93 |

**\*\*Categorical variables expressed as frequency and percentage; continuous variables as mean ± SD.**

Biomass fuel exposure was reported by 46.5% of participants, with a mean exposure duration of 1.62 ± 1.93 years. Chemical exposure was less common, reported by 27.9%. A family history of respiratory disease was present in 18.6% of respondents.

These findings suggest that while smoking and biomass fuel exposure were present, the majority were non-smokers with comparatively limited exposure histories.

Table 3 outlines participants’ clinical backgrounds. A previous history of tuberculosis was reported by only 7% of respondents. Oxygen therapy had been used by 8.5%.

**Table 3:**
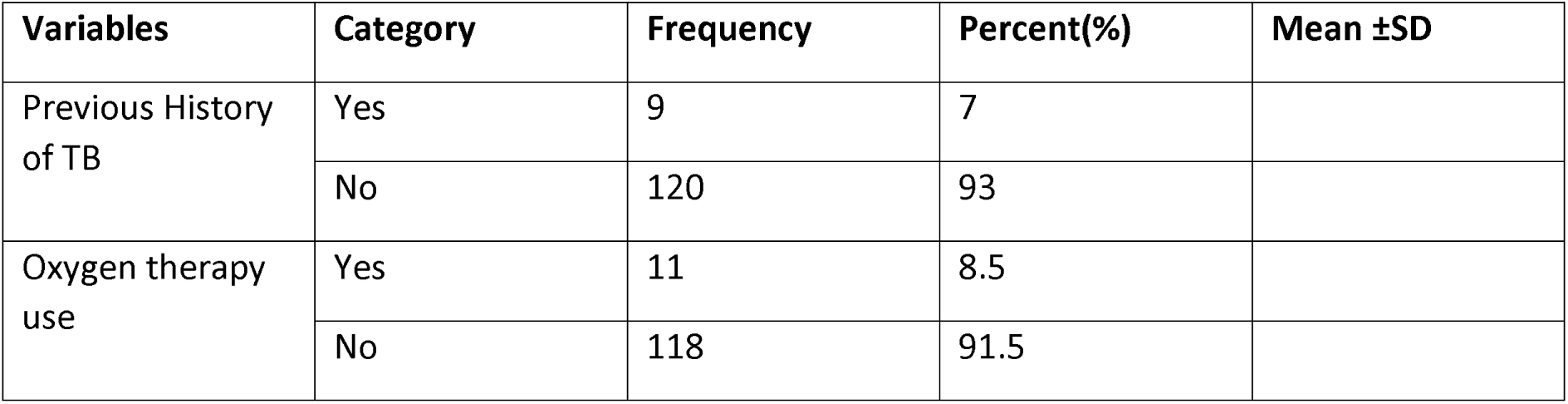
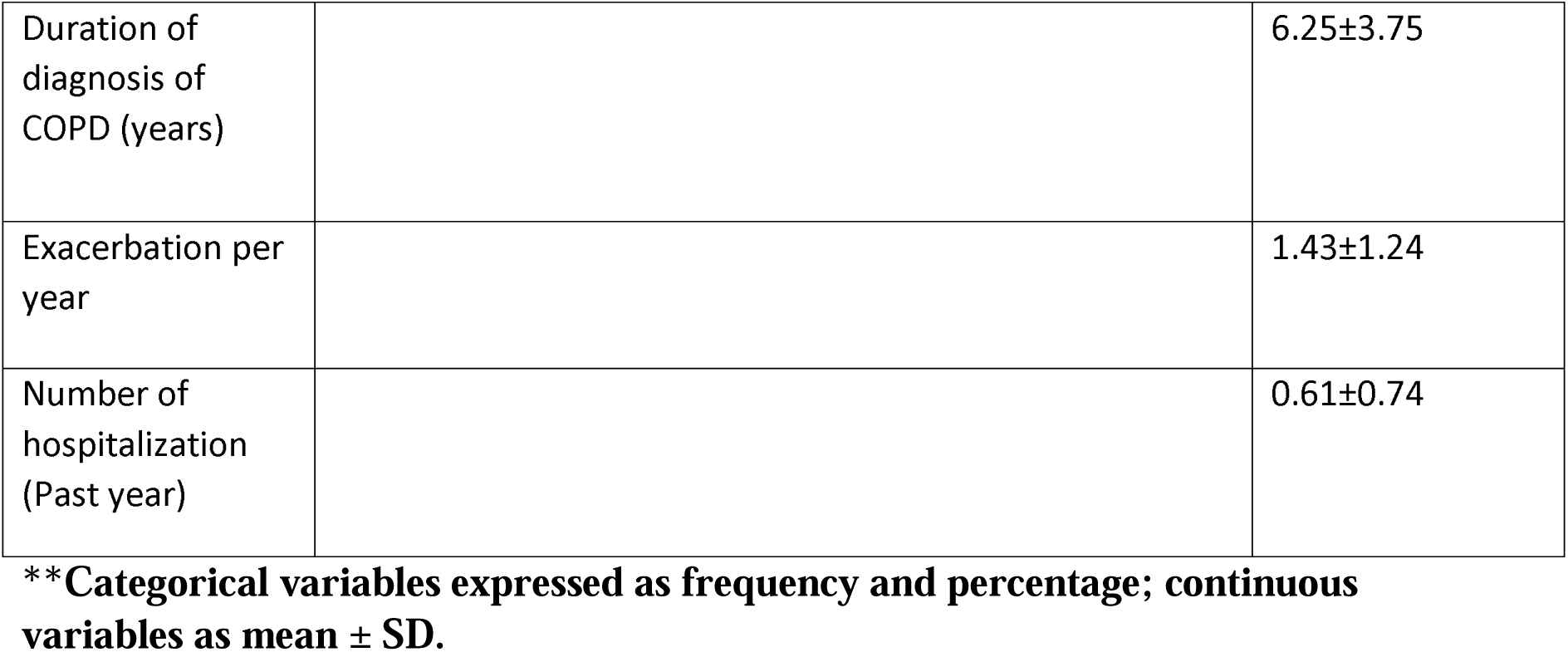
Distribution of respondents by clinical and disease-related variables (n = 129)

| Variables | Category | Frequency | Percent(%) | Mean ±SD |
| --- | --- | --- | --- | --- |
| Previous History of TB | Yes | 9 | 7 |  |
|  | No | 120 | 93 |  |
| Oxygen therapy use | Yes | 11 | 8.5 |  |
|  | No | 118 | 91.5 |  |

|  |  |  |
| --- | --- | --- |
| Duration of diagnosis of COPD (years) |  | 6.25±3.75 |
| Exacerbation per year |  | 1.43±1.24 |
| Number of hospitalization (Past year) |  | 0.61±0.74 |

**\*\*Categorical variables expressed as frequency and percentage; continuous variables as mean ± SD.**

The mean duration of COPD was 6.25 ± 3.75 years. Participants experienced an average of 1.43 ± 1.24 exacerbations per year. Hospital admissions were relatively infrequent, with a mean of 0.61 ± 0.74 admissions in the prior year.

Overall, respondents exhibited moderate disease duration with relatively low exacerbation and hospitalization frequencies

Table 4 presents a detailed overview of respiratory symptoms among the respondents, revealing a substantial symptomatic burden. Mucoid sputum was the most frequently reported type (61.2%), followed by purulent sputum (34.9%), while a smaller proportion (3.9%) experienced bloody sputum. Dyspnea was highly prevalent, with Grade 2 breathlessness—occurring while walking at a normal pace—being the most common (44.2%). Grade 3 and Grade 4 dyspnea affected 20.2% and 8.5% of participants respectively, whereas only 4.7% reported no dyspnea at all. Chronic cough was also widespread, reported by 72.1% of respondents, and hemoptysis was present in 5.4%.

**Table 4:**
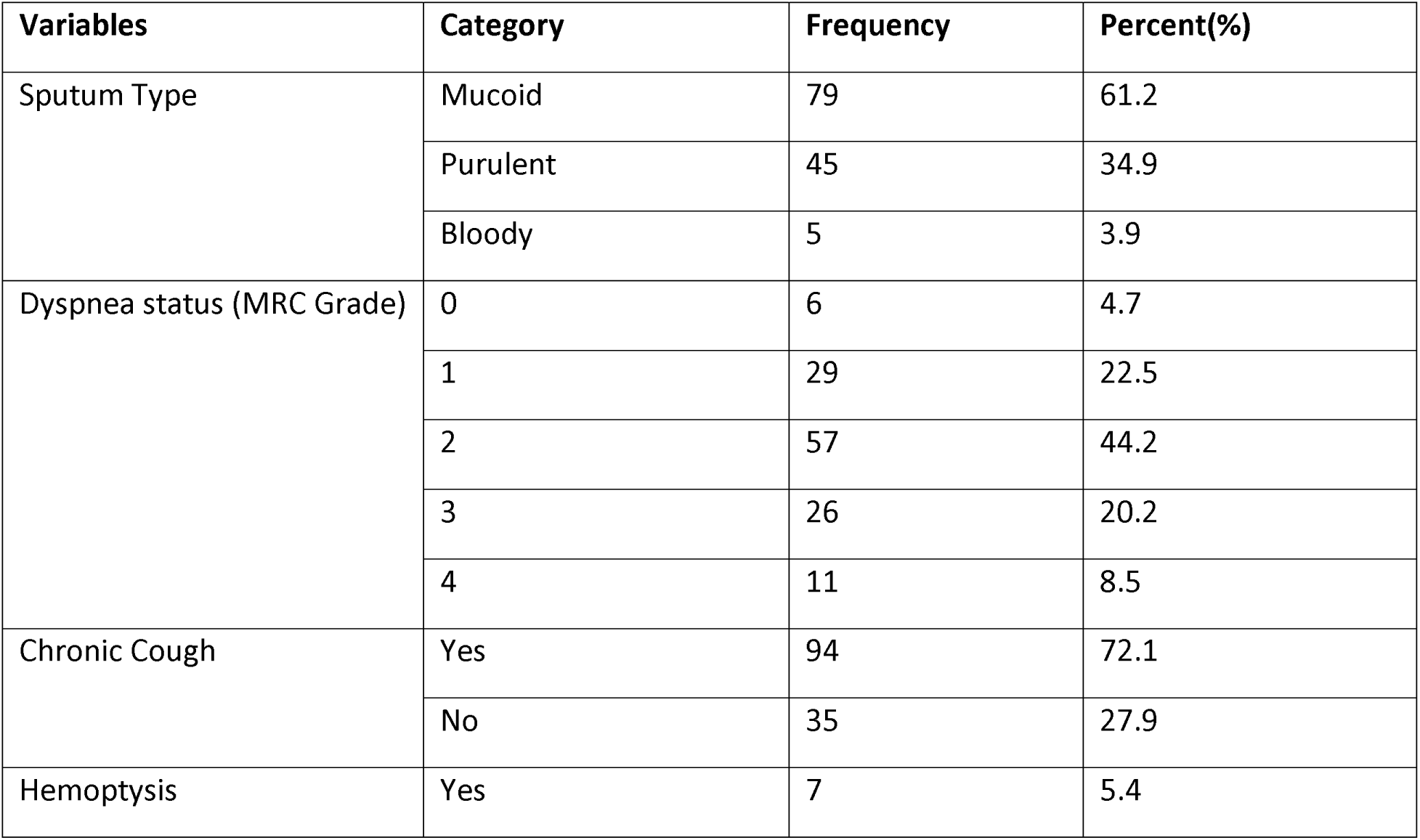
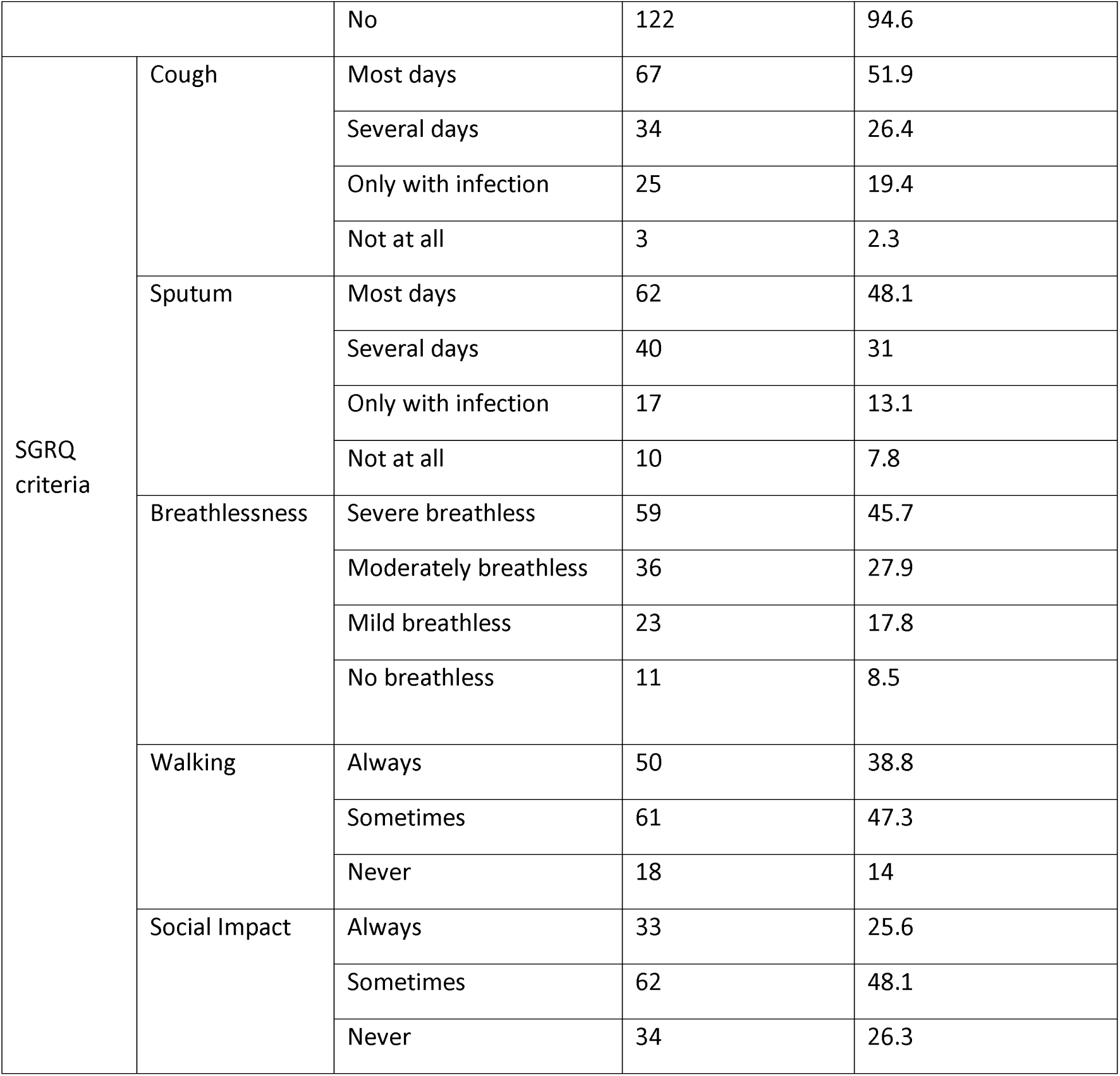
Distribution of respondents according to respiratory symptoms and functional status (n = 129)

| Variables | Category | Frequency | Percent(%) |
| --- | --- | --- | --- |
| Sputum Type | Mucoid | 79 | 61.2 |
|  | Purulent | 45 | 34.9 |
|  | Bloody | 5 | 3.9 |
| Dyspnea status (MRC Grade) | 0 | 6 | 4.7 |
|  | 1 | 29 | 22.5 |
|  | 2 | 57 | 44.2 |
|  | 3 | 26 | 20.2 |
|  | 4 | 11 | 8.5 |
| Chronic Cough | Yes | 94 | 72.1 |
|  | No | 35 | 27.9 |
| Hemoptysis | Yes | 7 | 5.4 |

|  |  |  |  |  |
| --- | --- | --- | --- | --- |
|  |  | No | 122 | 94.6 |
| SGRQ<br>criteria | Cough | Most days | 67 | 51.9 |
|  |  | Several days | 34 | 26.4 |
|  |  | Only with infection | 25 | 19.4 |
|  |  | Not at all | 3 | 2.3 |
|  | Sputum | Most days | 62 | 48.1 |
|  |  | Several days | 40 | 31 |
|  |  | Only with infection | 17 | 13.1 |
|  |  | Not at all | 10 | 7.8 |
|  | Breathlessness | Severe breathless | 59 | 45.7 |
|  |  | Moderately breathless | 36 | 27.9 |
|  |  | Mild breathless | 23 | 17.8 |
|  |  | No breathless | 11 | 8.5 |
|  | Walking | Always | 50 | 38.8 |
|  |  | Sometimes | 61 | 47.3 |
|  |  | Never | 18 | 14 |
|  | Social Impact | Always | 33 | 25.6 |
|  |  | Sometimes | 62 | 48.1 |
|  |  | Never | 34 | 26.3 |

Symptom patterns assessed using SGRQ criteria reflected considerable disease severity, as 51.9% experienced cough on most days and 48.1% had sputum production on most days, while 45.7% suffered from severe breathlessness. Functional limitations were notable, with 38.8% reporting that they were always limited in walking and 25.6% experiencing constant social interference due to their symptoms. Collectively, these findings underscore a marked respiratory and functional impairment among COPD patients, highlighting the extensive impact of the disease on daily life and social functioning.

Table 5 shows that 73.6% of respondents had bronchiectasis on imaging. Cylindrical bronchiectasis was most common (48.4%), followed by varicose (29.5%) and cystic (22.1%) types.

**Table 5:** Radiological features among study participants(n=129)

| Variables | Category | Frequency | Percent(%) |
| --- | --- | --- | --- |
| Bronchiectasis | Yes | 95 | 73.6 |
|  | No | 34 | 26.4 |
| Type of Bronchiectasis | Cylindrical | 46 | 48.4 |
|  | Varicose | 28 | 29.5 |
|  | Cystic | 21 | 22.1 |
| Lung Distribution | Unilateral | 16 | 12.4 |
|  | Bilateral | 26 | 20.2 |
|  | No involvement | 87 | 67.4 |
| Lobes Involved | Upper | 4 | 3.1 |
|  | Lower | 4 | 3.1 |
|  | Middle | 16 | 12.4 |
|  | Diffuse | 9 | 7 |
|  | None | 87 | 67.4 |
|  | Upper & Middle | 4 | 3.1 |
|  | Middle & Lower | 5 | 3.9 |
| Mucus Plugging | Yes | 24 | 18.6 |
|  | No | 105 | 81.4 |
| Emphysema | Yes | 45 | 34.9 |
|  | No | 84 | 65.1 |

Lobar involvement was variable, with the middle lobe affected most frequently (12.4%). Diffuse involvement (7%) and combined patterns (upper+middle or middle+lower) were also observed. Mucus plugging was present in 18.6%, and emphysema in 34.9%.

These findings indicate a substantial burden of structural lung abnormalities, with bronchiectasis being widespread and heterogeneous in distribution.

Table 6 describes the biochemical and pulmonary function characteristics of the respondents, showing notable variation across individuals. Serum albumin levels ranged from 2.99 to 4.72 g/dL, with a mean of 3.79 ± 0.41 g/dL, suggesting that most participants were within or slightly below normal nutritional and protein status. CRP levels ranged from 2.0 to 17.2 mg/L, with a mean of 3.89 ± 3.68 mg/L, indicating that while many respondents had low-grade inflammation, a subset exhibited markedly elevated inflammatory activity. Pulmonary function measurements demonstrated substantial heterogeneity; the mean FEV (% predicted) was 59.2 ± 17.02%, spanning from very severe obstruction to values approaching normal, and the mean FVC (% predicted) was 73.61 ± 16.94%, reflecting patterns consistent with both obstructive and mild restrictive components. Overall, these findings indicate moderate airflow limitation on average, with considerable physiological variability among the COPD patients assessed.

**Table 6:**
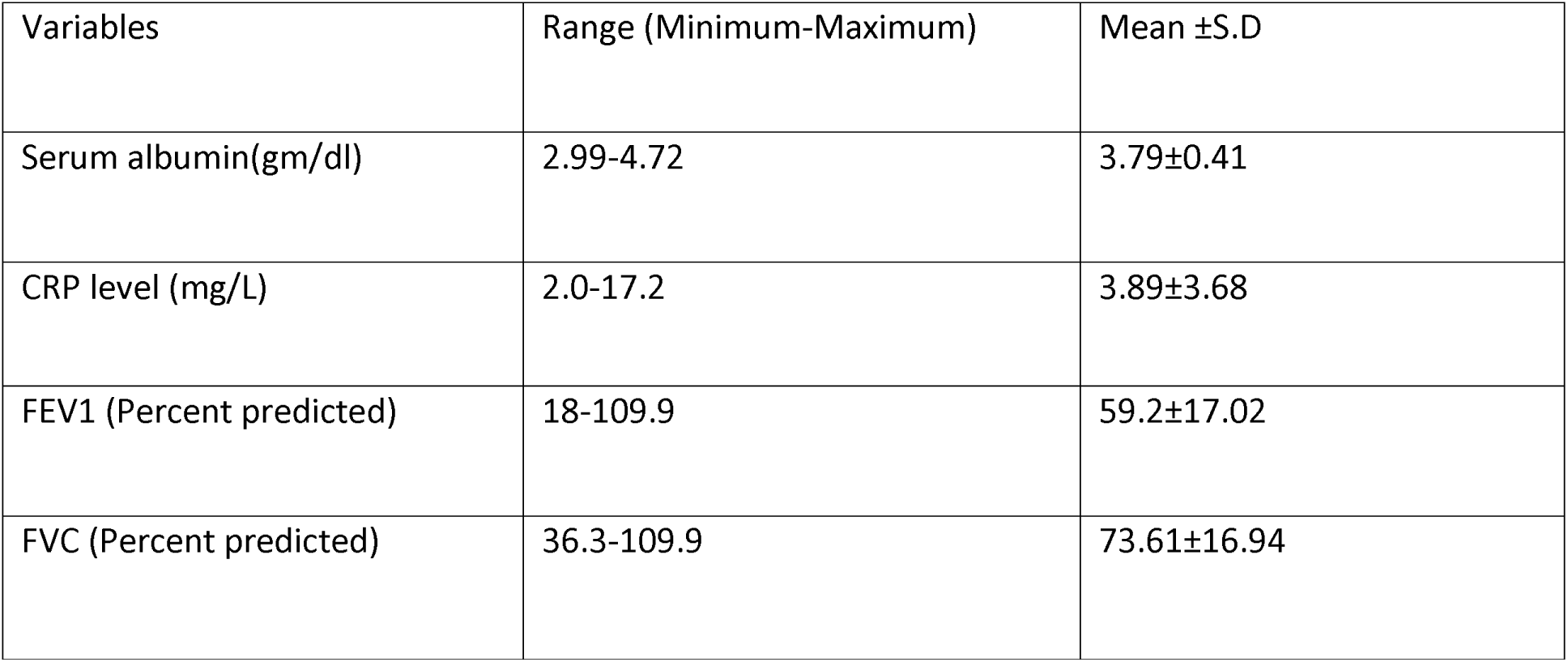
Laboratory and pulmonary function parameters of respondents (n=129)

Table 7 reveals that 40.3% of respondents reported poor quality of life, while 33.3% rated it as fair. Only 18.6% reported good quality of life, and 7.8% reported excellent quality.

**Table 7:**
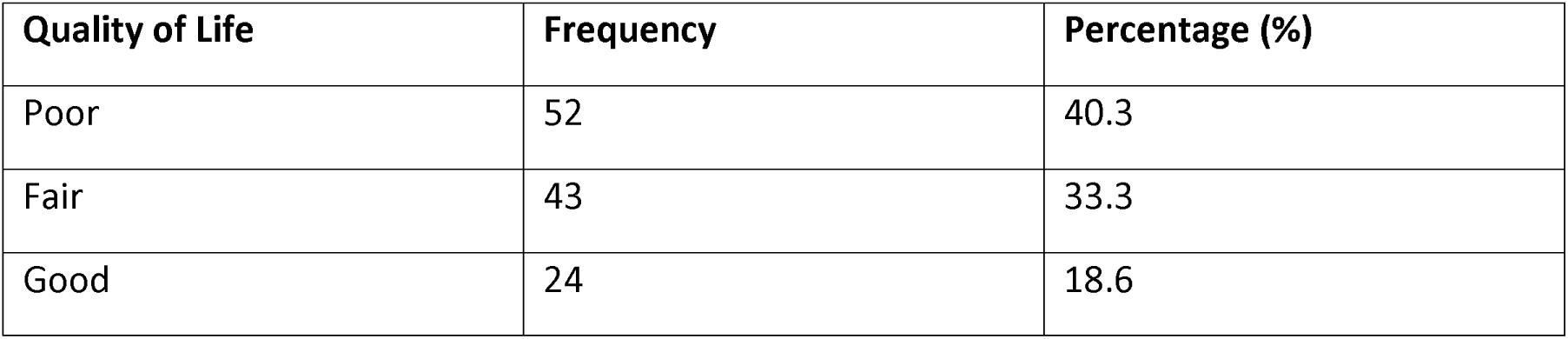
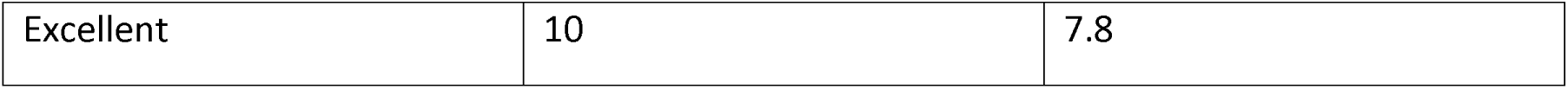
Distribution of respondents by quality of life score(n=129)

Thus, over 70% experienced fair-to-poor quality of life, reflecting substantial impairment due to their condition.

Table 8 shows a highly significant difference in COPD duration between groups. Participants with bronchiectasis had a mean COPD duration of 7.58 ± 3.36 years, compared to 2.51 ± 1.67 years among those without it (t = 8.418, p < 0.001).

**Table 8.**
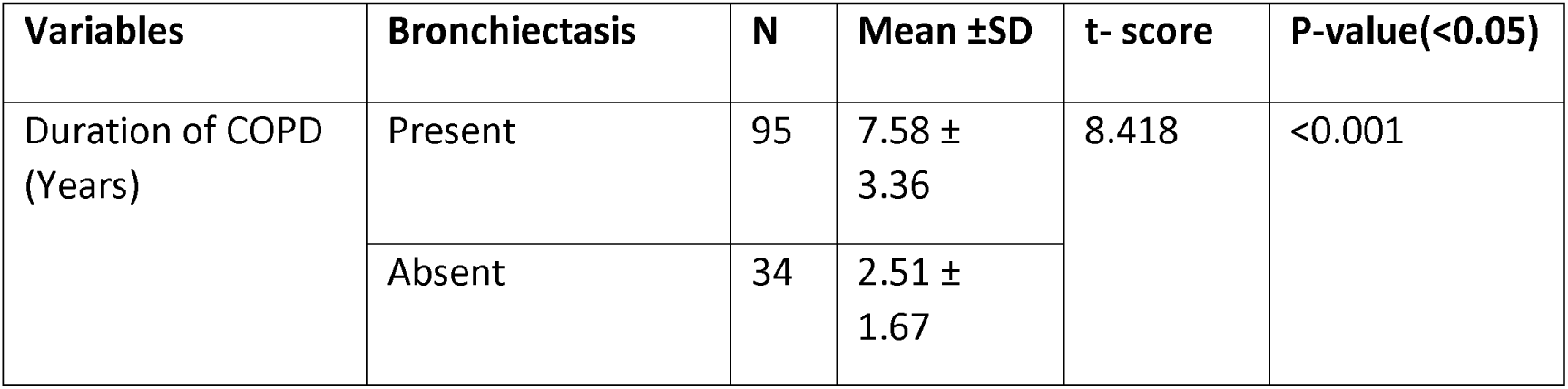
Association between duration of COPD and Bronchiectasis.

| Variables | Bronchiectasis | N | Mean ±SD | t- score | P-value(<0.05) |
| --- | --- | --- | --- | --- | --- |
| Duration of COPD (Years) | Present | 95 | 7.58 ± 3.36 | 8.418 | <0.001 |
| | Absent | 34 | 2.51 ± 1.67 | | |

This indicates that longer COPD duration is strongly associated with the development of bronchiectasis.

Table 9 highlights several important associations reflecting the multifactorial nature of bronchiectasis in COPD. Rural residents showed a markedly higher prevalence of bronchiectasis (91.9%) compared with those living in urban areas (66.3%), a difference that was statistically significant (χ^2^ = 8.901, p = 0.003). Smoking habit was also significantly associated with bronchiectasis (χ^2^ = 6.067, p = 0.048); however, the pattern was unconventional, as never-smokers demonstrated the highest prevalence (80.7%), suggesting that environmental and non-smoking factors may play a substantial role.

**Table 9.** Association between categorical variables and bronchiectasis.

| Variables | Category | Bronchiectasis |  | Chi-Square | P-value(<0.05) |
| --- | --- | --- | --- | --- | --- |
|  |  | Present | Absent |  |  |
| Residence | Urban | 61 (66.3%) | 31(33.7%) | 8.901 | 0.003 |
|  | Rural | 34 (91.9%) | 3(8.1%) |  |  |
| Smoking habit | Current | 21 (60%) | 14 (40%) | 6.067 | 0.048 |
|  | Former | 7 (63.6%) | 4 (36.4%) |  |  |
|  | Never | 67 (80.7%) | 16 (19.3%) |  |  |
| Gender | Male | 46 (64.8%) | 25 (35.2%) | 6.379 | 0.012 |
|  | Female | 49 (84.5%) | 9 (15.5%) |  |  |
| Mucus Plugging | Yes | 24 (25.3%) | 71 (74.7%) | 10.55 | 0.001 |
|  | No | 0** (0%) | 34 (100%) |  |  |
| Daily Cough | Most Days | 51 (76.1) | 16 (23.9) | 1.95 | 0.022 |
| Frequency |  |  |  |  |  |
|  | Several Days | 23 (67.6) | 11 (32.4) |  |  |
|  | Only with infection | 18 (72) | 7 (28) |  |  |
|  | Not at all | 3 (100) | 0** (0) |  |  |
| Social Interference | Very Often | 24 (72.7) | 9 (27.7) | 6.23 | 0.044 |
|  | Sometimes | 51 (82.3) | 11 (17.7) |  |  |
|  | Never | 20 (58.8) | 14 (41.2) |  |  |

**\*\* Fisher's exact test was done where cell count was less than 5**

Gender differences were pronounced, with females exhibiting a significantly higher prevalence (84.5%) than males (64.8%) (χ^2^ = 6.379, p = 0.012). Mucus plugging showed a perfect association—every participant with mucus plugging had bronchiectasis (χ^2^ = 10.55, p = 0.001), underscoring its strong diagnostic value. Daily cough frequency also emerged as a significant factor (χ^2^ = 1.95, p = 0.022), indicating that more frequent coughing was linked with a higher likelihood of bronchiectasis. Social interference due to symptoms showed a similar significant association (χ^2^ = 6.23, p = 0.044), reflecting how disease severity translates into functional limitations. Together, these findings emphasize that bronchiectasis in COPD is influenced by a complex interplay of demographic, behavioral, physiological, and symptomatic characteristics.

Table 10 shows a weak but significant negative correlation between smoking duration and FEV % predicted (r = −0.174, p = 0.048). Longer smoking history was associated with reduced lung function.

**Table 10.** Correlation between Duration of smoking and FEV1 (%)

| Variables | N | r | P value (<0.05) |
| --- | --- | --- | --- |
| Duration of smoking (years) ×<br>FEV<sub>1</sub> (% predicted) | 129 | -0.174 | 0.048 |

Table 11 demonstrates a weak but significant inverse correlation (r = −0.211, p = 0.016), indicating that higher inflammation corresponds to lower serum albumin levels.

**Table 11.**
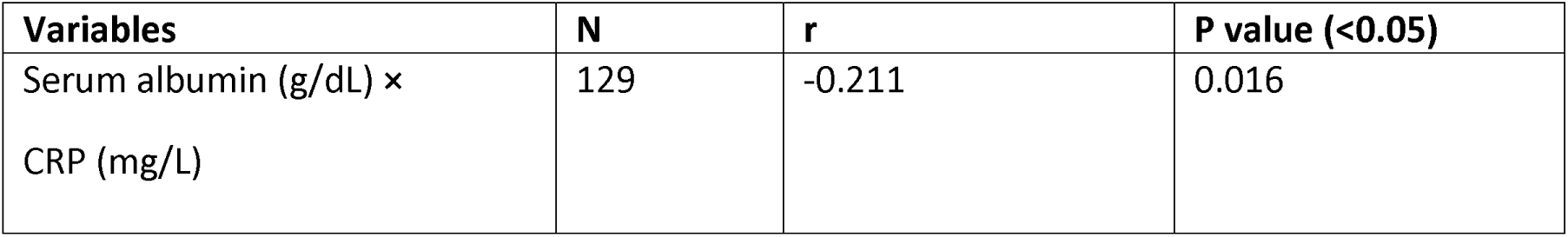
Correlation between Serum Albumin and CRP Levels among Respondents (n = 129)

Table 12 demonstrates a clear and strong association between the pattern of lung involvement and the presence of mucus plugging (χ^2^ = 61.948). None of the patients with no lung involvement showed mucus plugging, indicating a clean radiological pattern in this subgroup. In contrast, plugging was present in half of the individuals with unilateral involvement, suggesting that localized disease already predisposes to airway obstruction. The prevalence was even higher among those with bilateral involvement, where 61.5% exhibited mucus plugging, reflecting a more extensive and diffuse disease process. Overall, the findings show that as the extent of lung involvement increases—from none, to unilateral, to bilateral—the likelihood of mucus plugging rises substantially, reinforcing the link between structural lung damage and impaired airway clearance.

**Table 12.** Association between Lung Distribution and Mucus Plugging among Respondents (n = 129)

| Lung Distribution | Mucus Plugging |  | Chi-Square | P-value(<0.05) |
| --- | --- | --- | --- | --- |
|  | Yes | No |  |  |
| Unilateral | 8 (50%) | 8 (50%) | 61.948 | <0.001 |
| Bilateral | 16 (61.5%) | 10 (38.5%) |  |  |
| No Involvement | 0 (0%) | 87 (100%) |  |  |

The regression analysis identified two independent predictors of bronchiectasis, underscoring the influence of both environmental and behavioral factors. Residence emerged as a significant determinant; individuals living in rural areas had markedly higher odds of developing bronchiectasis compared with urban residents (AOR = 5.82, p = 0.019), suggesting that rural environmental exposures, delayed healthcare access, or biomass fuel use may contribute substantially to disease risk. Smoking habit also remained an independent predictor, with current smokers showing significantly higher odds of bronchiectasis than non-smokers (AOR = 3.69, p = 0.049). Together, these findings indicate that even after adjusting for other factors, place of residence and smoking behavior play key roles in shaping bronchiectasis risk among COPD patients.

**Table 13:** Binary Logistic Regression Analysis of Factors Associated with Bronchiectasis.

| Predictor Variables | p- value | Adjusted Odds ratio | 95% CI<br>(Confidence Interval) |
| --- | --- | --- | --- |
| Residence | 0.019 | 5.82 | [1.34 – 25.29] |
| Smoking Habit | 0.049 | 3.69 | [1.01 – 13.49] |

## Discussion

The strikingly high prevalence of bronchiectasis in this cohort—73.6% of COPD patients—emphasizes that structural airway damage is far more than an incidental finding in Bangladeshi COPD populations. This rate is at the upper end of the globally reported range, which varies significantly across studies depending on diagnostic techniques, patient selection, and disease severity; systematic reviews and meta-analyses have reported prevalence ranging from 25% to nearly 70% in COPD cohorts using high-resolution computed tomography (HRCT) for diagnosis^1,2^. Such a high burden suggests that in resource-limited settings like Bangladesh, where repeated infections, environmental exposures, and limited access to preventive therapies are common, bronchiectatic changes may accumulate more readily and progress more aggressively than in better-resourced populations.

Clinically, this cohort presented with a heavy symptom burden: a large majority reported chronic cough (72.1%) and sputum production, including purulent (34.9%) and bloody (3.9%) sputum, while hemoptysis was seen in 5.4%. Dyspnea was nearly universal, with 44.2% of patients at MRC grade 2, 20.2% at grade 3, and 8.5% at grade 4. These findings broadly reflect the typical “overlap phenotype,” where patients with COPD and bronchiectasis suffer from persistent sputum and increased exacerbation risk^3,4^. Indeed, previous work has shown that bronchiectasis in COPD is associated with more frequent exacerbations, greater inflammation, and microbial colonization^3,4^, and our data reinforce this: the mean exacerbation rate of 1.43 per year and average hospitalization rate of 0.61 per year indicate a clinically unstable population.

Radiologically, cylindrical bronchiectasis was by far the most common subtype (48.4%), followed by varicose (29.5%) and cystic (22.1%) forms. These proportions are consistent with patterns reported in earlier COPD–bronchiectasis cohorts, in which cylindrical disease often predominates, possibly because it represents an earlier or less severe form of bronchial dilatation^1,5^. The lung distribution findings, with 12.4% of patients manifesting unilateral disease and 20.2% bilateral involvement, further suggest a heterogeneous structural burden in this population. Importantly, mucus plugging was present in only 18.6% of participants, but critically, all patients with mucus plugging had bronchiectasis; none without plugging did. This complete co-occurrence underscores the pathophysiological importance of mucus retention in the development or maintenance of structural lung damage, aligning with other studies that highlight plugging as a marker of more severe or advanced bronchiectasis^3,6^.

One of the most notable divergences from many Western or high-income country cohorts lies in the risk factor and demographic profile. In our logistic regression, residence emerged as a strong independent predictor: rural participants had nearly six fold increased odds of bronchiectasis (OR 5.82; 95% CI 1.34–25.29), a finding that is not frequently reported in studies from developed settings. While urban air pollution is often blamed for higher respiratory disease burden, our data suggest that in Bangladesh, rural populations may face heightened risk of structural lung damage, possibly due to biomass fuel exposure, limited access to healthcare, repeated untreated infections, or delayed diagnosis. Indeed, nearly half of our participants (46.5%) reported exposure to biomass fuel, pointing toward non-smoking sources of airway injury commonly prevalent in rural, low-resource settings.

Interestingly, smoking habit was also significantly associated with bronchiectasis (OR 3.69; 95% CI 1.01–13.49). However, on cross-tabulation, the highest proportion of bronchiectasis was observed among those who never smoked (80.7%), followed by former (63.6%) and current smokers (60%). This somewhat counterintuitive distribution suggests that in our population, non-smoking exposures (like biomass) or recurrent infections may play a more dominant role than smoking in the pathogenesis of bronchiectasis. This diverges from many studies in Western cohorts where smoking is more directly implicated in both COPD and bronchiectasis overlap. Such a pattern underscores the distinct epidemiological landscape in South Asia, where risk factors like indoor air pollution, poverty, and infection persist outside the classical paradigm of tobacco-driven lung disease^8^.

Gender differences also arose in our cohort: female participants had a higher prevalence of bronchiectasis (84.5%) compared to males (64.8%, p = 0.012). This gender disparity could relate to socio-environmental exposures: women in rural Bangladesh may bear a disproportionate burden of biomass exposure, cooking-related particulate inhalation, and household pollutants. Biological susceptibility or healthcare-seeking behavior may also contribute; previous literature suggests that hormonal and immunological differences can influence airway inflammation and repair, though these mechanisms remain incompletely understood in overlap populations.

From a systemic perspective, the correlation analyses revealed a weak but statistically significant inverse relationship between CRP and serum albumin (r = −0.211, p = 0.016), indicating that a higher inflammatory burden is modestly but meaningfully associated with lower nutritional status. This aligns with emerging data in non-cystic fibrosis bronchiectasis, where hypoalbuminemia (as a negative acute-phase reactant) correlates with worse disease severity^10^. In our cohort, the mean albumin was 3.79 g/dL, suggesting that while frank malnutrition may not be predominant, low-grade systemic inflammation may subtly erode protein reserves. This finding has important clinical implications: in a resource-limited setting, where advanced biomarkers or sophisticated monitoring may be sparse, albumin could serve as an accessible proxy to gauge systemic disease burden and guide therapeutic interventions.

The functional impairment indicated by spirometry was considerable: mean FEV was 59.2 % predicted, and mean FVC was 73.6 % predicted. There was a weak but statistically significant negative correlation between smoking duration and FEV (r = − 0.174, p = 0.048), reinforcing the cumulative deleterious effects of tobacco on lung function, albeit modest in this cohort. While smoking remains a well-established cause of airflow limitation in COPD, our findings suggest that in this population, non-tobacco insults—such as biomass exposure or infection—may compound structural damage, thereby contributing to reduced airflow independent of smoking history.

Quality of life, as measured by SGRQ, was severely compromised: 40.3% of patients rated their health-related quality of life as “poor,” 33.3% as “fair,” only 18.6% as “good,” and a mere 7.8% as “excellent.” Such a distribution indicates that over 70% of the cohort lives with substantial functional, psychological, and social impairment. Notably, bronchiectasis was significantly associated with greater social interference (p = 0.044), suggesting that the overlap phenotype exacts a pronounced human cost beyond spirometric decline—it affects daily life, social roles, and psychological well-being. This aligns with regional data showing that COPD severely diminishes quality of life in Bangladeshi populations^11^, but our findings extend this by illustrating how structural lung disease may amplify that burden even further.

The strong association between COPD duration and bronchiectasis (mean duration 7.58 ± 3.36 years in overlap vs. 2.51 ± 1.67 years in non-bronchiectasis, p < 0.001) underscores the progressive nature of this overlap syndrome. This temporal relationship aligns with the pathophysiological concept that persistent airway inflammation, impaired mucociliary clearance, and recurrent infections progressively remodel the bronchial architecture leading to irreversible dilatation^6^. Over time, these processes likely reinforce each other, establishing a vicious cycle of inflammation, infection, and structural damage, consistent with the chronic airway disease model described in prior literature^6^.

Comparatively, many epidemiological studies in high-income countries emphasize colonization with *Pseudomonas aeruginosa* as a driver of exacerbations and decline in overlap patients^3,4^. While our study did not assess microbial colonization, the clinical profile—frequent exacerbations, persistent purulent sputum, and radiological bronchiectasis—strongly suggests that bacterial infection likely plays a critical role. Given the resource constraints in Bangladesh, limited access to sputum culture or molecular diagnostics may hamper early pathogen identification, but our data make a compelling case for integrating microbiological surveillance (where feasible) into standard care protocols, especially for those with frequent symptoms or exacerbations.

Therapeutically, the high prevalence and deep clinical burden of bronchiectasis in Bangladeshi COPD patients carry important implications. Standard COPD management, which typically emphasizes bronchodilators and inhaled corticosteroids, may be insufficient or even suboptimal for patients with coexisting bronchiectasis. In bronchiectasis, therapies such as airway clearance techniques, long-term macrolide antibiotics, and targeted antimicrobial regimens have shown benefit in reducing exacerbations and improving quality of life^12^. However, these therapies are not yet routinely integrated into COPD treatment algorithms in many low- and middle-income countries. Our findings suggest that clinicians in Bangladesh should maintain a high clinical suspicion of bronchiectasis in COPD patients—especially those with long disease duration, persistent sputum, frequent exacerbations, or radiographic abnormalities—and consider early referral for HRCT and adjunctive management.

Furthermore, the inverse correlation between albumin and CRP suggests that simple, widely available serum biomarkers may play a valuable role in risk stratification and monitoring, particularly where advanced diagnostics are limited. Maintaining or improving nutritional status could be a pragmatic intervention to reduce systemic inflammation, potentially attenuating disease progression or improving resilience to exacerbations.

From a public health and policy perspective, our study highlights several under-recognized priorities. First, the rural–urban disparity in bronchiectasis risk suggests environmental and healthcare access inequities that need to be addressed. Public health strategies aimed at reducing biomass exposure, improving indoor air quality, and strengthening primary respiratory care in rural areas may mitigate some of the root causes of structural lung disease. Second, training for healthcare providers on the overlap phenotype and its management should be prioritized. Without recognition, many COPD patients with bronchiectasis may remain undertreated, with persistent symptoms, poor quality of life, and repeated hospitalizations.

While the cross-sectional design of this study limits causal inference, the robust associations between disease duration, symptoms, radiology, inflammation, and nutritional markers provide a strong foundation for future longitudinal research. Prospective studies are needed to determine whether early detection of bronchiectasis in COPD patients, followed by targeted therapy, can reduce exacerbation frequency, slow lung function decline, and improve patient-centered outcomes in this setting. Microbiological studies, including sputum culture and molecular techniques, would help elucidate the pathogen spectrum in this population and guide antibiotic stewardship. Finally, cost-effectiveness analyses of HRCT screening strategies, airway clearance interventions, and biomarker-based monitoring are warranted to guide resource allocation in low-resource healthcare systems.

In conclusion, our findings shed light on a highly prevalent, deeply burdensome COPD–bronchiectasis overlap phenotype in Bangladesh. Many of the clinical features echo those found in global studies, but our data also reveal unique regional patterns—particularly the role of non-smoking exposures, rural residence, and systemic nutritional-inflammation interplay—that challenge conventional paradigms and call for locally adapted diagnostic and therapeutic strategies. Addressing these challenges through heightened clinical vigilance, targeted therapy, and public health interventions could transform care for a large and vulnerable population of COPD patients, improving both structural outcomes and quality of life.

## Conclusion

This study reveals an alarmingly high burden of bronchiectasis among COPD patients in Bangladesh, with 73.6% of the cohort showing radiologically confirmed bronchiectatic changes. The overlap phenotype was associated with heavier symptom load, more functional limitation, and poorer overall health status. Patients with bronchiectasis had longer COPD duration, more persistent cough, more sputum production (including purulent), and greater social interference—confirming a more severe clinical trajectory than COPD alone.

Radiologically, cylindrical bronchiectasis predominated, and mucus plugging showed a perfect association with bronchiectasis, underscoring its diagnostic importance. Bilateral involvement was notably linked to mucus plugging, reflecting deeper structural compromise in advanced disease.

Several key determinants emerged. Rural residence and smoking were strong, independent predictors of bronchiectasis. Yet paradoxically, non-smokers still carried a high disease burden, highlighting the substantial contribution of biomass exposure, infections, and environmental pollutants common in the Bangladeshi context. Females also had a disproportionately higher prevalence, likely tied to household biomass exposure patterns.

Systemic inflammation was evident through the significant inverse relationship between CRP and serum albumin, reinforcing the systemic nature of the disease. Lung function impairment was moderate but heterogeneous, and quality of life was markedly reduced, with more than 70% reporting fair-to-poor HRQoL.

Overall, this study demonstrates that bronchiectasis is not a minor comorbidity but a major structural complication of COPD in Bangladesh. The findings highlight the need for earlier detection, better risk-factor mitigation, and a shift toward phenotype-specific management to reduce exacerbations, improve lung function, and enhance quality of life.

## Recommendations

Based on the findings of this study, it is recommended that routine HRCT screening be incorporated for COPD patients who present with persistent sputum, frequent exacerbations, or longer disease duration, as early identification of bronchiectasis can significantly improve management outcomes. Treatment strategies should be adapted to include phenotype-specific care such as airway clearance techniques, long-term macrolides, and culture-directed antibiotics rather than relying solely on standard COPD protocols. Public health interventions targeting smoking cessation and reduction of biomass exposure—particularly among rural populations and women—are crucial to mitigating preventable risk factors. Strengthening provider awareness at the primary care level is essential for earlier referral of high-risk patients, while simple biomarkers like CRP and serum albumin should be routinely monitored to assess systemic inflammation and guide nutritional support. Expanding access to pulmonary rehabilitation and improving rural respiratory services can help reduce functional impairment and quality-of-life deficits. Finally, longitudinal research and microbiological surveillance are warranted to better understand disease progression, guide antibiotic stewardship, and support development of cost-effective diagnostic and therapeutic policies tailored to the Bangladeshi context.

## Data Availability

The data supporting this study are not publicly available due to ethical reasons but are available from the corresponding author upon reasonable request.

